# Exploring the relationship between schizophrenia and cardiovascular disease: A genetic correlation and multivariable Mendelian randomization study

**DOI:** 10.1101/2021.07.09.21260260

**Authors:** Rada R Veeneman, Jentien M Vermeulen, Abdel Abdellaoui, Eleanor Sanderson, Robyn E Wootton, Rafik Tadros, Connie R Bezzina, Damiaan Denys, Marcus R Munafò, Karin JH Verweij, Jorien L Treur

## Abstract

**Importance:** Individuals with schizophrenia have a reduced life-expectancy compared to the general population, largely due to an increased risk of cardiovascular disease (CVD). Clinical and epidemiological studies have been unable to fully unravel the nature of this relationship.

**Objective:** Investigate genetic correlations and potential bi-directional effects between liability to schizophrenia and CVD.

**Design, setting, and participants:** We obtained summary-data of genome-wide-association studies of schizophrenia (N=130,644), heart failure (N=977,323), coronary artery disease (N=332,477), systolic and diastolic blood pressure (N=757,601), heart rate variability (N=46,952), QT interval (N=103,331), early repolarization and dilated cardiomyopathy ECG patterns (N=63,700). We computed genetic correlations with linkage disequilibrium score regression and conducted bi-directional Mendelian randomization (MR). With multivariable MR, we investigated whether associations were mediated by smoking, body mass index, physical activity, lipid levels, or type 2 diabetes. To ensure robustness, we applied a range of sensitivity methods.

**Main outcomes and measures:** Schizophrenia, heart failure, coronary artery disease, systolic blood pressure, diastolic blood pressure, heart rate variability, QT interval, early repolarization, dilated cardiomyopathy.

**Results:** Genetic correlations between liability to schizophrenia and CVD were close to zero (−0.02 to 0.04). With MR, we found robust evidence that liability to schizophrenia increases heart failure risk. This effect remained consistent with multivariable MR. There was also evidence that liability to schizophrenia increases early repolarization risk, largely mediated by BMI and lipid levels. Finally, there was evidence that liability to schizophrenia increases heart rate variability, a direction of effect contrasting previous studies. In the other direction, there was weak evidence that higher systolic, but not diastolic, blood pressure increases schizophrenia risk.

**Conclusions and relevance:** Our findings indicate that liability to schizophrenia increases the risk of heart failure, and that this is not mediated by key health behaviours. This is consistent with the notion that schizophrenia is characterised by a systemic dysregulation of the body (including inflammation and oxidative stress) with detrimental effects on the heart. To decrease cardiovascular mortality among schizophrenia patients, priority should lie with optimal treatment and interventions in early stages of psychoses. We also identified early repolarization, currently understudied, as a potential CVD marker among patients with schizophrenia.

## Introduction

Schizophrenia is a serious mental disorder affecting up to 1% of the population.^1^ The life-expectancy of individuals diagnosed with schizophrenia is approximately 15–20 years shorter than that of the general population^2^ – a major reason being cardiovascular mortality.^3^ This is reflected by the increased prevalence of cardiovascular diseases (CVD), including coronary artery disease and heart failure, and risk factors, including high blood pressure and abnormal electro-cardiogram (ECG) patterns, amongst individuals with schizophrenia.^4–8^

There are broadly two potential, not mutually exclusive, explanations for this co-morbidity. First, there may be a shared aetiology. Low birth weight, pre-term birth and maternal malnutrition during pregnancy are associated with an increased risk of both schizophrenia and CVD in offspring.^9,10^ There is also evidence for shared genetic influences, but only for a limited number of cardio-metabolic traits and using considerably smaller samples than currently available.^11^ Second, there may be causal effects. The predominant hypothesis is that schizophrenia increases the risk of CVD. Schizophrenia is characterised by elevated cortisol levels, dysfunction of the autonomic nervous system, inflammation, lipid abnormalities, oxidative stress and increased platelet reactivity^4,9,10^, all of which contribute to the development and progression of CVD.^12^ Reverse causal effects have also been proposed, with markers of CVD preceding and potentially inducing psychosis.^10,13^

While systemic characteristics of schizophrenia may lead to CVD, there are also potential mediators. Anti-psychotic medication use, which can cause central obesity, hypertension, and abnormal lipid patterns^14^, may in turn increase CVD risk.^15^ This doesn’t explain all excess cardiovascular mortality, as patients who do not use anti-psychotics are also at increased risk of CVD.^5,14^ Other potential mediators are smoking, poor diet and lack of physical activity, all common in individuals with schizophrenia and risk factors for CVD.^16,17^ While conducting a randomized controlled trial (RCT) is not feasible, a powerful alternative is Mendelian Randomization (MR).^18^ MR mimics an RCT by using genetic variants as proxies, or ‘instrumental variables’, for the proposed risk factor.^19^ Because genetic variants are randomly passed on from parents to offspring, bias from confounders can be circumvented (provided core assumptions are met).^28^

We capitalize on the availability of large genetic samples and sophisticated methods to elucidate the nature of the relationship between schizophrenia and CVD. Using summary-level data of genome-wide association studies (GWAS), we: (1) compute genetic correlations to determine genome-wide overlap between schizophrenia and CVD risk, (2) perform univariable MR to test if liability to schizophrenia increases CVD risk, (3) perform univariable MR to test if liability to CVD increases schizophrenia risk, and (4) perform multivariable MR to test if key health behaviours mediate associations of liability to schizophrenia on CVD risk.

## Methods

The analysis plan was pre-registered at https://osf.io/fprew.

### Data

For schizophrenia, we obtained European-based summary statistics from the largest available GWAS^20^. For CVD, we selected eight phenotypes often linked to schizophrenia, for which sufficiently large GWAS were available (extensive justification in **supplement**). These entailed two *clinical endpoints*; coronary artery disease^21^ and heart failure^22^, and six *markers of CVD risk*; systolic blood pressure^23^, diastolic blood pressure^23^, heart rate variability (HRV)^24^, QT interval^25^, early repolarization ECG pattern^26^, and dilated cardiomyopathy ECG pattern^26^ (see **Table 1** for phenotype definitions).

**Table 1.** Overview of the main phenotypes, details on their reference and how they were measured, the number of SNPs included in the MR instruments and the genetic correlations between schizophrenia and the cardiovascular disease phenotypes

| Phenotype | GWAS reference | How the phenotype was measured | Sample size | n SNPs in MR instrument | Genetic correlation with schizophrenia |
| --- | --- | --- | --- | --- | --- |
| Schizophrenia | Ripke *et al*, 2020 | Cases were individuals diagnosed with a schizophrenia spectrum disorder, based on DSM-IV criteria | 53,386 cases<br>77,258 controls | 185 | - |
| Coronary artery disease | Nelson *et al*, 2017 | Cases were individuals with a fatal or nonfatal myocardial infarction, percutaneous transluminal coronary angioplasty (PTCA), coronary artery bypass grafting (CABG), chronic ischemic heart disease, or angina | 71,602 cases<br>260,875 controls | 56 | $rg=-0.02$, $SE=0.03$, $p=0.439$ |
| Heart failure | Shah *et al*, 2020 | Cases were individuals with a clinical diagnosis of heart failure of any aetiology (no inclusion criteria based on left ventricle ejection fraction) | 47,309 cases<br>930,014 controls | 12 | $rg=-0.01$, $SE=0.04$, $p=0.696$ |
| Systolic blood pressure | Evangelou *et al*, 2018 | Mean of two systolic blood pressure measurements (if available), manual or automatic (or both). Measured in millimetres of mercury (mmHg) | 757,601 | 237 | $rg=-0.01$, $SE=0.02$, $p=0.699$ |
| Diastolic blood pressure | Evangelou *et al*, 2018 | Mean of two diastolic blood pressure measurements (if available), manual or automatic (or both). Measured in millimetres of mercury (mmHg) | 757,601 | 158 | $rg=0.001$, $SE=0.02$, $p=0.965$ |
| Heart rate variability (HRV) | Nolte *et al*, 2017 | The root mean square of the successive differences of inter beat intervals (RMSSD), which reflects heart rate variability | 46,952 MR exposure sample<br>26,523 MR outcome sample + *rg* analysis | 9 | $rg=-0.01$, $SE=0.05$, $p=0.850$ |
| QT interval | Arking *et al*, 2014 | The time from the start of the Q wave to the end of the T wave as read from an ECG. Individual were excluded if there was atrial fibrillation, atrial flutter, presence of QRS duration >120 msec or presence of left/right bundle branch block | 103,331 | 68 | $rg=0.04$, $SE=0.03$, $p=0.142$ |
| Early repolarization ECG-pattern | Verweij *et al*, 2020 | The amplitude at <u>+44 msec</u> (after the R wave) of the electrocardiogram (ECG), which coincided with the early repolarization criteria. The genome-wide significant SNPs were followed up in an independent cohort (Lifelines) to confirm their association with early repolarization diagnosis (1,253 cases, 11,463 controls) | 63,700 | 10 | $rg=-0.02$, $SE=0.04$, $p=0.600$ |
| Dilated cardiomyopathy ECG-pattern | Verweij *et al*, 2020 | The amplitude at <u>-18 msec</u> (before the R wave) of the electrocardiogram (ECG), which showed strong overlap with dilated cardiomyopathy risk. The genome-wide significant SNPs were followed up in an independent cohort (UK biobank) to confirm their association with dilated cardiomyopathy diagnosis (1,375 cases, 241,325 controls) | 63,700 | 29 | $rg=0.004$, $SE=0.04$, $p=0.892$ |

For multivariable MR, we selected five potential mediators of effects of liability to schizophrenia on CVD. These capture health behaviours, or their downstream consequences, which are particularly prevalent among individuals with schizophrenia: smoking (initiation^27^ and lifetime smoking^28^), body mass index (BMI)^29^, physical activity^30^, lipid levels (total cholesterol and triglycerides^31^– may be increased as a result of anti-psychotic medication), and type-2-diabetes^32^.

For MR, sample overlap between some of the exposure and outcome variables was prevented by excluding overlapping UK-Biobank participants. For further explanation of the samples see Supplement.

### Genetic correlations

Genetic correlations were computed using linkage disequilibrium score (LDSC) regression.^33^ The genetic correlation is based on the estimated slope from the regression of the product of z-scores from two GWAS on the LD score and represents the covariation between two traits based on all polygenic effects captured by the included SNPs. We filtered GWAS summary statistics to only include the 1,290,028 million SNPs from the HapMap 3 European reference panel, used to provide the genome-wide LD information.^33,34^

### Mendelian randomization

We conducted bi-directional MR to assess evidence for effects of (genetic) liability to schizophrenia on CVD risk, and vice versa, of (genetic) liability to CVD on schizophrenia risk. MR relies on three assumptions: (1) the genetic variants robustly associate with the exposure; (2) the genetic variants are independent of confounders; (3) the genetic variants do not affect the outcome, except through their effect on the exposure (Figure 1A). ‘Horizontal pleiotropy’, a variant associating with multiple traits, may violate assumptions 2 and 3, if the variant associates with the outcome directly or via a confounding factor.^18^

**Figure 1.**
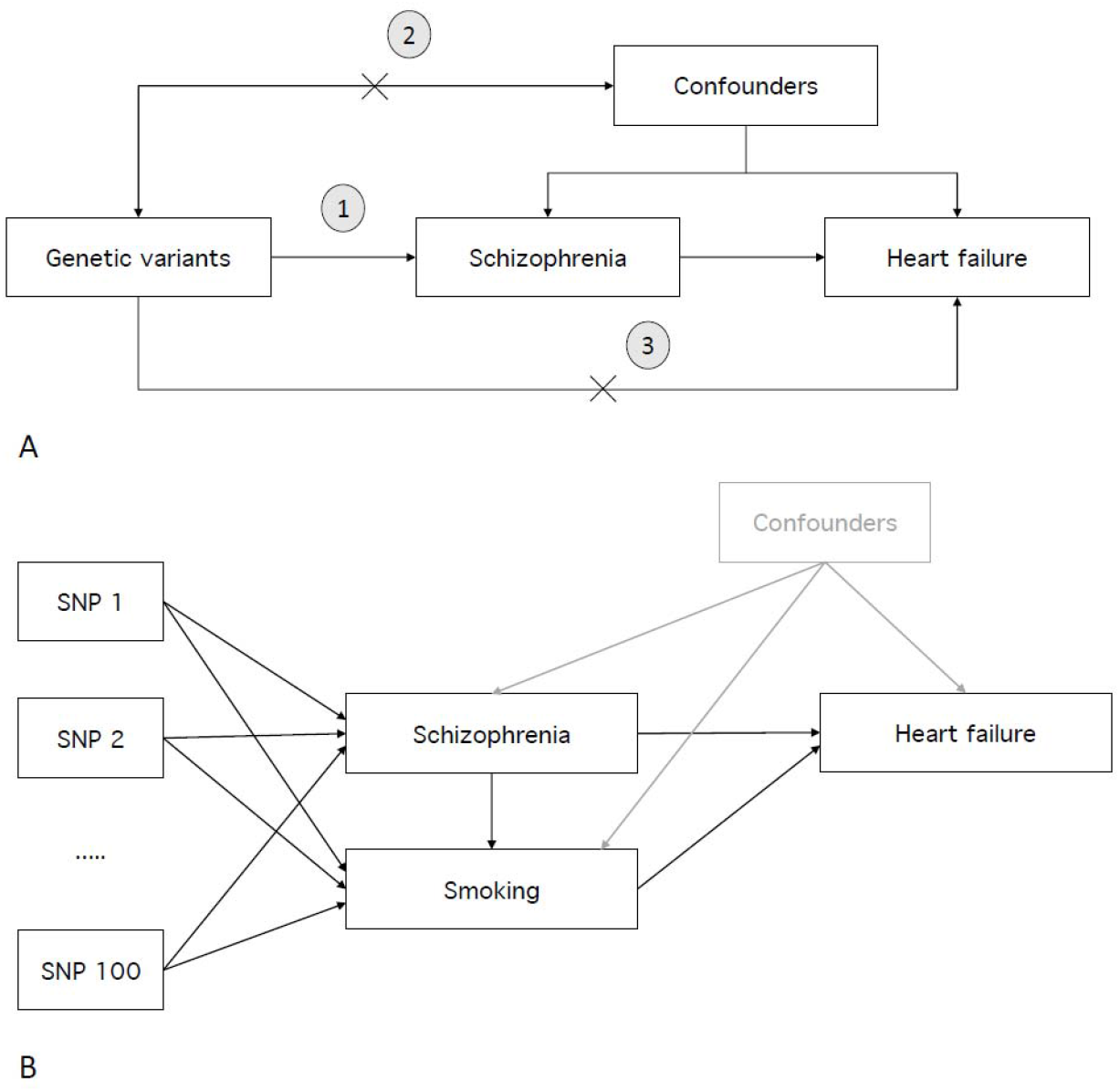
The Mendelian randomization (MR) principle. A) The validity of MR relies on three assumptions, the genetic variants used in the instrument must (1) associated robustly with the exposure variable (e.g. schizophrenia), (2) be independent of confounders, and (3) not directly affect the outcome variable (e.g. heart failure), except through their effect on the exposure. B) Multivariable MR allows the inclusion of an additional variable, besides the main exposure variable. In the current study we aimed to test whether key health behaviours (e.g. smoking) mediate part of the effect of schizophrenia on cardiovascular disease (risk). In this example, if the inclusion of smoking results in a (considerable) decrease in the direct effect of schizophrenia on heart failure, that implies that smoking mediates some of that relationship.

The main method was inverse-variance weighted (IVW) regression. Independent SNPs that reached genome-wide significance (*p*<5E-08) in the exposure (e.g., schizophrenia) GWAS were extracted to form instrumental variables. SNP-outcome effects were obtained from the outcome (e.g., heart failure) GWAS. A ratio estimate was obtained by dividing the effect a SNP has on the outcome by the effect it has on the exposure. Individual SNP-effects were weighted by the inverse of their variance and estimates of all SNPs combined into one estimate. IVW provides the first indication of whether the exposure impacts the outcome, assuming all assumptions are met. To verify its validity, we applied a range of sensitivity methods less vulnerable to bias: weighted median regression^35^, weighted mode regression^36^, MR-Egger regression^37^, MR-PRESSO (Mendelian Randomization Pleiotropy RESidual Sum and Outlier)^38^, GSMR (Generalised Summary-data-based Mendelian Randomisation)^39^, and Steiger filtering^40^. We also performed leave-one-out analyses, repeating IVW analyses after removing each SNP one at a time, and computed Cochran’s Q to assess heterogeneity between the SNP-estimates in each instrument. For further details see **Supplement**.

Using multivariable MR, health behaviours were added to see whether associations between liability to schizophrenia as the exposure and CVD as the outcome diminished, which could indicate mediation (**Figure 1B**).^41^ For each univariable analysis we added each health behaviour separately (we did not combine them to prevent violation of the linearity and homogeneity assumptions^42^). We applied multivariable MR-Egger to evaluate robustness (**Supplement**).

A finding was considered robust when results were consistent across methods. Because sensitivity methods rely on stricter assumptions than IVW, they are less powerful. Their statistical evidence, but not effect size, will therefore be weaker, even for a true effect. We describe findings as showing no clear evidence, weak evidence, evidence or strong evidence, taking into account IVW and the sensitivity methods, adhering to the broad interpretation of p-values described by Sterne and Davey Smith (2001).^43^

Analyses were performed in R version 3.6.3, using packages: ‘TwoSampleMR’, ‘GSMR’, ‘psych’, and ‘MR-PRESSO’.

## Results

### Genetic correlations

Genetic correlations, reflecting overlap between genome-wide liability to schizophrenia and CVD, were all close to zero (**Table 1**).

### Univariable Mendelian randomization

Instruments for schizophrenia and CVD were sufficiently strong (F-statistic>10; **Table S1**). Bi-directional univariable MR results are depicted in **Table 2**.

**Table 2.**
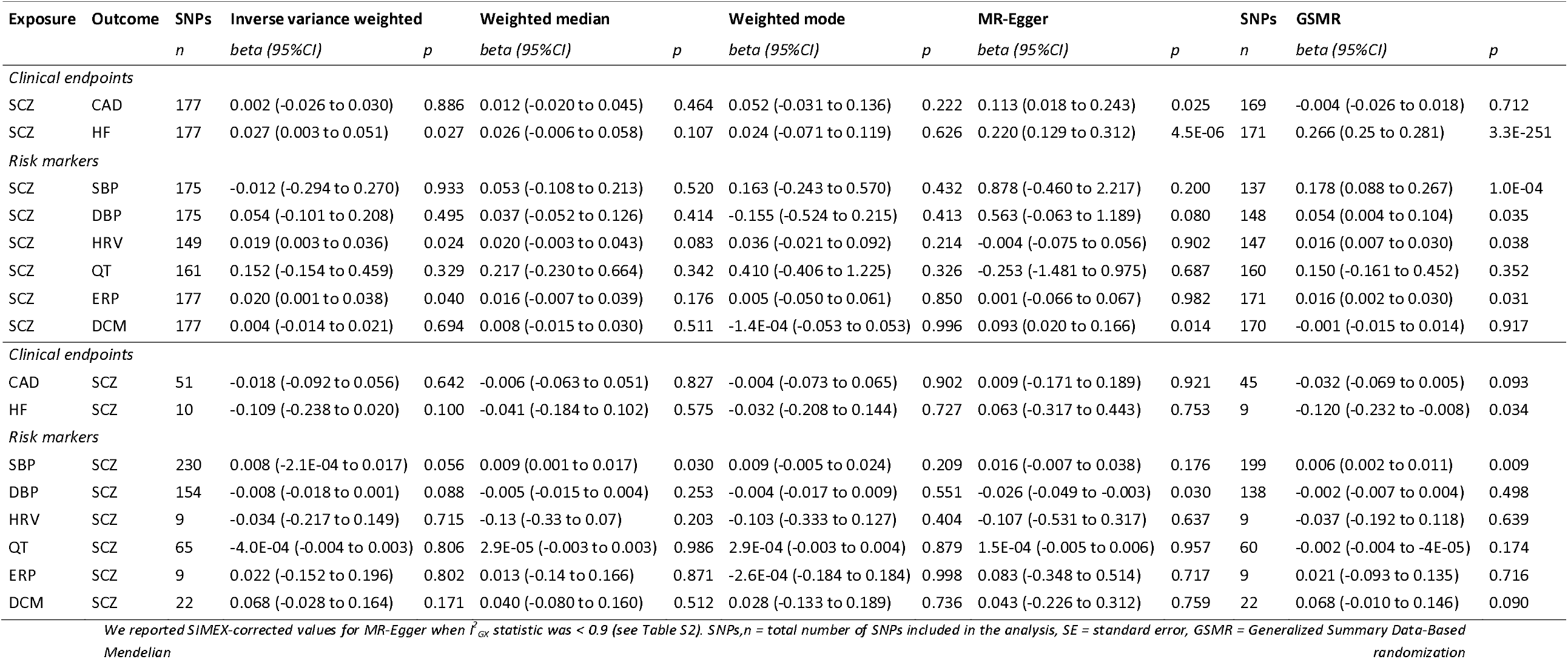
Results of univariable, bi-directional Mendelian randomization analyses between liability to schizophrenia (SCZ) and coronary artery disease (CAD), systolic blood pressure (SBP), diastolic blood pressure (DBP), heart failure (HF), heart rate variability (HRV), QT interval (QT), early repolarization ECG pattern (ERP), and dilated cardiomyopathy ECG pattern (DCM)

### Liability to schizophrenia on CVD

There was evidence that liability to schizophrenia increases heart failure risk (beta_IVW_=0.027, 95% Confidence intervals=0.003 to 0.051, p=0.027). Weighted median and weighted mode regression confirmed this with consistent effect sizes, but weaker statistical evidence. GSMR showed much stronger evidence for a positive effect, of greater magnitude (beta=0.266, CIs=0.25 to 0.281, p=3.3E-251). While the MR-Egger intercept showed strong evidence for horizontal pleiotropy (−0.013, CIs=-0.019 to −0.007, p=9.4E-05; **Table S2**), its slope also indicated strong evidence for a positive effect (beta=0.220, CIs=0.129 to 0.312, p=4.5E-06). There was strong evidence for heterogeneity across SNPs (Cochran’s Q p=2.3E-04; **Table S3**). MR-PRESSO detected one outlier, but elimination of this SNP did not have a large impact (**Table S4**). Steiger filtering did not find SNPs that explained more variance in the outcome than the exposure, suggesting no reverse causality (**Table S5**). While leave-one-out analysis showed SNP rs13107325 to have a relatively large impact, the effect size and statistical evidence remained considerable after removal (**Figure S1**).

There was evidence that liability to schizophrenia increases early repolarization pattern (beta_IVW_=0.020, CIs=0.001 to 0.038, p=0.040). While weighted median and GSMR confirmed this, weighted mode and MR-Egger regression did not (beta=0.005, CIs=-0.050 to 0.061, p=0.850, and beta=0.001, CIs=-0.066 to 0.067, p=0.982, respectively). There was strong evidence for heterogeneity (p=6.8E-09), but the MR-Egger intercept indicated that this was not due to horizontal pleiotropy (p=0.580). MR-PRESSO detected one outlier, which did not impact the results. Steiger filtering excluded one SNP resulting in a slightly weaker effect.

There was evidence that liability to schizophrenia increases HRV (beta_IVW_=0.019, CIs=2.5E-03 to 0.036, p=0.024). Weighted median, weighted mode and GSMR were consistent, while MR-Egger was not. There was evidence for heterogeneity (p=0.009), but this was likely not due to horizontal pleiotropy (MR-Egger intercept p=0.499). Excluding a single outlier with MR-PRESSO did not impact the results, while excluding six SNPs with Steiger filtering slightly attenuated the effect.

No other analyses showed clear evidence for association (**Table 2**).

### Liability to CVD on schizophrenia risk

There was weak evidence that increased systolic blood pressure increases schizophrenia risk (beta_IVW_=0.008, CIs=-2.1E-04 to 0.017, p=0.056). All sensitivity methods confirmed this, with the strongest evidence from GSMR (beta=0.006, CIs=0.002 to 0.011, p=0.009). There was strong evidence for heterogeneity (p=4.6E-68), but the MR-Egger intercept indicated that this was not due to horizontal pleiotropy (p=0.491). MR-PRESSO detected 14 outliers, yet elimination of these SNPs did not change the results. Steiger filtering removed 16 SNPs, resulting in even stronger evidence (beta_IVW_=0.010, CIs=0.003 to 0.016, p=0.006). Leave-one-out analysis showed that removing SNP rs11191548 considerably decreased the effect size and weakened statistical evidence (**Figure S2**). There was very weak evidence that liability to increased diastolic blood pressure decreases schizophrenia risk (beta_IVW_=-0.008, CIs=-0.018 to 0.001, p=0.088), but this was not corroborated by the sensitivity analyses.

No other analyses showed clear evidence for association (**Table 2**).

### Multivariable Mendelian randomization

For relationships that showed evidence of association, multivariable results are shown in **Figure 2**. All other multivariable results are in **Tables S6-S22**. The conditional F statistic indicated sufficient instrument strength, except for physical activity which ranged between ∼8 and ∼11 (**Table S15**).^44^

**Figure 2.**
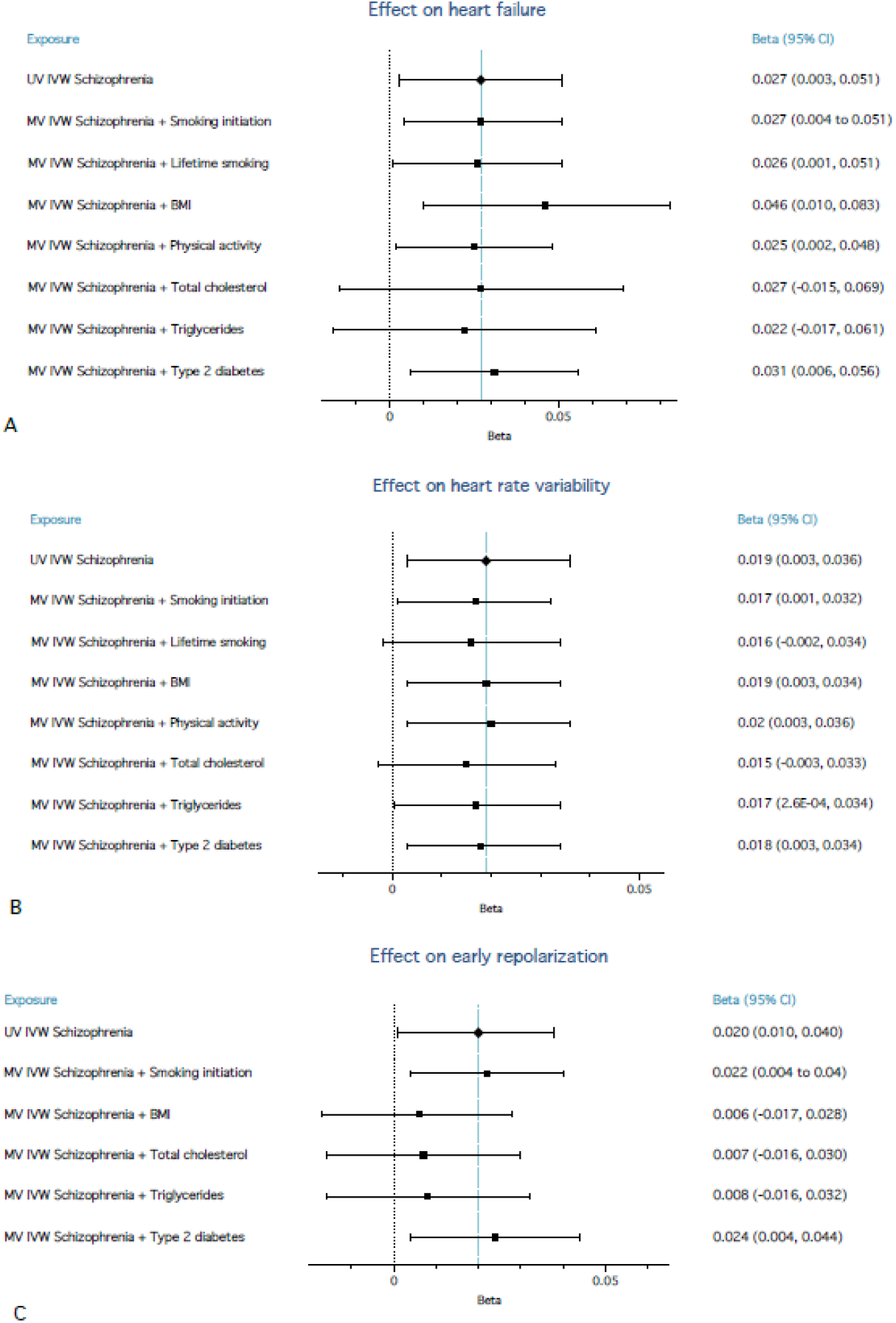
Forest plots of multivariable Mendelian randomization (MR) analyses from liability to schizophrenia on heart failure (A), liability to schizophrenia on heart rate variability (B), and liability to schizophrenia on early repolarization (C), showing the direct effect of liability to schizophrenia on the respective outcomes. Each of the health behaviours were added in separate multivariable MR analyses. Note that lifetime smoking and physical activity could not be added for the analyses with early repolarization as the outcome (C), because there was considerable sample overlap between the GWAS these were based on and the GWAS for early repolarization (overlap in UK-Biobank participants).

Most of the health behaviours had a larger impact on heart failure than did liability to schizophrenia. When added in multivariable MR, only BMI had a noteworthy impact on the direct effect of liability to schizophrenia on heart failure. BMI increased the direct effect of liability to schizophrenia by ∼70% (beta_IVW_=0.046, CIs=0.01 to 0.083, p=0.013). MR-Egger confirmed the effect size and direction (beta=0.030, CIs=-0.033 to 0.093, p=0.354). The MR-Egger intercept indicated no horizontal pleiotropy (p=0.537; **Table S16**). BMI itself showed a positive association with heart failure (beta_IVW_=0.447, CIs=0.34 to 0.555, p=3.4E-16), suggesting it is negatively associated with schizophrenia. We conducted post-hoc analyses with underweight (versus normal weight) and overweight (versus normal weight) as potential mediators (**Supplement**). Adding underweight did not change the main effect, while overweight increased it by ∼30% (conditional F-statistic for underweight was low (2.28)).

The effect of liability to schizophrenia on HRV remained consistent after adding health behaviours.

The effect of liability to schizophrenia on early repolarization pattern decreased when adding BMI, total cholesterol and triglycerides, with 70% (beta_IVW_=0.006, CIs=-0.017 to 0.028, p=0.617), 65% (beta_IVW_=0.007, CIs=-0.017 to 0.031, p=0.549), and 60% (beta_IVW_=0.008, CIs=-0.016 to 0.032, p=0.538), respectively. MR-Egger results were consistent and the intercepts did not indicate horizontal pleiotropy.

## Discussion

We computed genome-wide genetic correlations between schizophrenia and eight important CVD phenotypes, all of which were nearly zero. Using MR, there was evidence that liability to schizophrenia increases heart failure risk. This effect was not mediated by health behaviours in multivariable MR. There was evidence that liability to schizophrenia increases early repolarization pattern, largely mediated by BMI and lipid levels. There was also evidence that liability to schizophrenia increases HRV, a direction of effect contrasting clinical observations. Finally, there was weak evidence that increased systolic blood pressure increases schizophrenia risk, which was not confirmed with diastolic blood pressure.

The lack of evidence for genetic correlation between schizophrenia and CVD is striking, given previously reported phenotypic correlations.^3^ In contrast, considerable positive genetic correlations between schizophrenia and immune-mediated diseases mirrored earlier epidemiological evidence.^45^ Our findings indicate that there is minimal shared genetic aetiology, implying that phenotypic associations between schizophrenia and cardiovascular disease are due to causal effects or confounding. It should be noted that our analyses test the effects of liability to schizophrenia, and not of a schizophrenia diagnosis *per se*. It may be that an actual diagnosis of schizophrenia, and all detrimental factors associated with it, drive associations with cardiovascular disease, and we were not able to capture that.

With MR, we found evidence that liability to schizophrenia increases CVD risk. MR employs a selection of genetic variants, as opposed to the genome-wide approach for genetic correlations. MR might therefore find an association that was ‘cancelled out’ by opposing effects in a genome-wide correlation.^46^ There was robust evidence that liability to schizophrenia increases heart failure risk, confirming the high prevalence of heart failure among individuals with schizophrenia.^47^ MR’s powerful premise and the robustness across sensitivity analyses, allows us to say with more certainty that this is due to causal effects. Important to note is that the MR-Egger intercept indicated horizontal pleiotropy, such that genetic variants for schizophrenia also exerted some effect on heart failure, independent of schizophrenia. The effect on heart failure was not mediated by health behaviours, which is particularly noteworthy for smoking. Smoking rates among individuals with schizophrenia are remarkably high and smoking increases cardiovascular mortality.^17^ Our findings are in line with the notion that schizophrenia is characterized by a systemic dysregulation of the body, including inflammation and oxidative stress, which promotes cardiac alterations and ultimately heart failure.^9,48^ This implies that changing health behaviours – while useful to improve overall health – is not sufficient to reduce cardiovascular mortality among patients with schizophrenia. To prevent heart failure, priority should lie with optimally treating and interventions in early stages of psychosis, thereby resolving detrimental systemic effects.

There was evidence that liability to schizophrenia increases early repolarization pattern, corroborating reports that early repolarization disproportionately affects patients with schizophrenia.^49,50^ Historically, this pattern was considered a normal phenotype^51^, but recent evidence linked it to an increased risk of sudden cardiac death.^52^ Sudden cardiac death is particularly prevalent in patients with schizophrenia, making early repolarization a potentially important risk marker. The association with early repolarization pattern declined when correcting for BMI, total cholesterol, and triglycerides. This suggests that among individuals with schizophrenia this pattern can be improved by lowering BMI and lipid levels. This provides new leads to elucidate the, currently poorly understood, aetiology of early repolarization.^52^

Liability to schizophrenia was associated with higher HRV, while patients with schizophrenia present with lower HRV in the clinic – partly due to antipsychotic medication use.^53,54^ This discrepancy may be explained by the fact that we made use of two separate samples (one GWAS for schizophrenia, a separate GWAS for HRV). The number of individuals with a schizophrenia diagnosis – and thus antipsychotic use – was likely low in the HRV GWAS. Another important consideration is that the HRV GWAS data were not corrected for heart rate, which correlates strongly with HRV and is usually higher in individuals with schizophrenia^8^. Finally, lower HRV may result from the systemic presentation of schizophrenia, which our measure of liability did not capture. Future studies tracking HRV before and after schizophrenia is diagnosed would help clarify these contradicting findings.

Finally, there was evidence that higher systolic, but not diastolic, blood pressure increases schizophrenia risk. This corroborates a large cohort study reporting that, in men, higher blood pressure in adolescence predicts schizophrenia later in life.^55^ Higher blood pressure has also been reported in individuals who were at risk of, but had not yet developed, psychosis.^56^ Combined, this suggests dysfunction in the autonomic nervous system precedes the onset of schizophrenia. A potentially important confounder is smoking, as it affects both blood pressure^57^ and schizophrenia^58^. However, in a post-hoc multivariable MR analysis, we found no change in effect when adding smoking (**Table S17**).

### Strengths and limitations

Our study has some important strengths. We were able to conduct powerful analyses, investigating disorders with a low prevalence on the population level (especially schizophrenia). We applied a wide range of rigorous MR sensitivity methods, which increases the robustness of our inferences, including multivariable MR which allowed us to investigate important mediators. Combined, this has kept the risk of bias from horizontal pleiotropy and reverse causality to a minimum.

There are also limitations to consider. Schizophrenia is a severe illness and those who suffer most may not have been able or willing to participate in research, causing selection bias.^59,60^ For early repolarization and dilated cardiomyopathy, it should be taken into consideration that these do not reflect the whole pattern, but rather a particular amplitude of the ECG that corresponds with the beginning of the respective patterns. For some relationships, there may have been temporality issues. Schizophrenia often arises in early adulthood, whereas heart failure and coronary artery disease develop later in life. Testing heart failure and coronary artery disease as exposures for schizophrenia is therefore imperfect. However, genetic risk to CVD can already have an impact early in life and therefore causal relationships are plausible.^61^ Finally, assortative mating, dynastic effects (‘genetic nurture’) and residual population stratification could not be accounted for. Future within-family Mendelian randomization analyses may be able to reduce such bias.^62^

### Clinical implications

We showed that a shared genetic aetiology is not the most likely mechanism underlying associations between schizophrenia and CVD. There was, however, evidence that liability to schizophrenia increases the risk of heart failure and early repolarization pattern. While the association with early repolarization pattern was largely mediated by BMI and lipid levels, the association with heart failure remained stable after adding key health behaviours. This implies that effective treatment and intervention in early psychosis is important to decrease excess cardiovascular mortality. Tracking of physical health and screening for CVD is currently done less frequently than advised in clinical guidelines, and start of treatment is often delayed.^63^ More thorough screening throughout psychiatric treatment must become a priority, in order to decrease the stark mortality gap between schizophrenia patients and individuals from the general population.

## Data Availability

All analyses in this study were based on publicly available (summary-level) GWAS data

## Acknowledgements

JLT is supported by a Young Investigator (NARSAD) Grant from the Brain & Behaviour Research Foundation. Specifically, the current work was made possible by the *Evelyn Toll Family Foundation*, whom we would like to greatly acknowledge for their support. KJHV, AA, and JLT are also supported by the Foundation Volksbond Rotterdam. AA is supported by ZonMw grant 849200011 from The Netherlands Organisation for Health Research and Development. We acknowledge SURFsara for the usage of the Lisa cluster computer (supported by NWO, 15725). CRB was supported by the Dutch Heart Foundation Netherlands Cardiovascular Research Initiative (CVON; PREDICT2 2018-30). MRM is a member of the MRC Integrative Epidemiology Unit at the University of Bristol (MC_UU_00011/7), and the National Institute for Health Research (NIHR) Biomedical Research Centre at the University Hospitals Bristol National Health Service (NHS) Foundation Trust, and the University of Bristol. REW is supported by a postdoctoral fellowship from the South-Eastern Regional Health Authority (2020024).

## Notes

### Competing Interest Statement

The authors have declared no competing interest.

### Author Declarations

Not relevant as these analyses were based on anonymous, summary-level data of large-scale public datasets

## References

1. Stępnicki P, Kondej M, Kaczor AA. Current concepts and treatments of schizophrenia. Molecules. 2018;23(8). doi:10.3390/molecules23082087

2. Hjorthøj C, Stürup AE, McGrath JJ, Nordentoft M. Years of potential life lost and life expectancy in schizophrenia: a systematic review and meta-analysis. The Lancet Psychiatry. 2017;4(4):295–301. doi:10.1016/S2215-0366(17)30078-0

3. Ringen PA, Engh JA, Birkenaes AB, Dieset I, Andreassen OA. Increased mortality in schizophrenia due to cardiovascular disease - a non-systematic review of epidemiology, possible causes and interventions. Front Psychiatry. 2014;5(SEP). doi:10.3389/fpsyt.2014.00137

4. De Hert M, Detraux J, Vancampfort D. The intriguing relationship between coronary heart disease and mental disorders. Dialogues Clin Neurosci. 2018;20(1):31–40. http://www.ncbi.nlm.nih.gov/pubmed/29946209. Accessed February 5, 2019.

5. Blom MT, Cohen D, Seldenrijk A, et al. Brugada Syndrome ECG Is Highly Prevalent in Schizophrenia. Circ Arrhythmia Electrophysiol. 2014;7(3):384–391. doi:10.1161/CIRCEP.113.000927

6. Koponen H, Alaräisänen A, Saari K, et al. Schizophrenia and sudden cardiac death—A review. Nord J Psychiatry. 2008;62(5):342–345. doi:10.1080/08039480801959323

7. Mitchell AJ, Vancampfort D, De Herdt A, Yu W, De Hert M. Is the prevalence of metabolic syndrome and metabolic abnormalities increased in early schizophrenia? a comparative meta-analysis of first episode, untreated and treated patients. Schizophr Bull. 2013;39(2):295–305. doi:10.1093/schbul/sbs082

8. Cohen H, Loewenthal U, Matar M, Kotler M. Association of autonomic dysfunction and clozapine: Heart rate variability and risk for sudden death in patients with schizophrenia on long-term psychotropic medication. Br J Psychiatry. 2001;179(2):167–171. doi:10.1192/BJP.179.2.167

9. Pillinger T, Enrico D’ambrosio •, Mccutcheon R, Howes OD. Is psychosis a multisystem disorder? A meta-review of central nervous system, immune, cardiometabolic, and endocrine alterations in first-episode psychosis and perspective on potential models. Mol Psychiatry. doi:10.1038/s41380-018-0058-9

10. Dieset I, Andreassen OA, Haukvik UK. Somatic Comorbidity in Schizophrenia: Some Possible Biological Mechanisms Across the Life Span. Schizophr Bull. 2016;42(6):1316–1319. doi:10.1093/schbul/sbw028

11. So HC, Chau KL, Ao FK, Mo CH, Sham PC. Exploring shared genetic bases and causal relationships of schizophrenia and bipolar disorder with 28 cardiovascular and metabolic traits. Psychol Med. 2019;49(8):1286–1298. doi:10.1017/S0033291718001812

12. Golia E, Limongelli G, Natale F, et al. Inflammation and cardiovascular disease: From pathogenesis to therapeutic target. Curr Atheroscler Rep. 2014;16(9). doi:10.1007/s11883-014-0435-z

13. Chen YL, Pan CH, Chang CK, et al. Physical illnesses before diagnosed as schizophrenia: A nationwide case-control study. Schizophr Bull. 2020;46(4):785–794. doi:10.1093/schbul/sbaa009

14. Vermeulen J, van Rooijen G, Doedens P, Numminen E, van Tricht M, de Haan L. Antipsychotic medication and long-term mortality risk in patients with schizophrenia; a systematic review and meta-analysis. Psychol Med. 2017;47(13):2217–2228. doi:10.1017/S0033291717000873

15. De Hert M, Detraux J, Van Winkel R, Yu W, Correll CU. Metabolic and cardiovascular adverse effects associated with antipsychotic drugs. Nat Rev Endocrinol. 2012;8(2):114–126. doi:10.1038/nrendo.2011.156

16. Bly MJ, Taylor SF, Dalack G, et al. Metabolic syndrome in bipolar disorder and schizophrenia: Dietary and lifestyle factors compared to the general population. Bipolar Disord. 2014;16(3):277–288. doi:10.1111/bdi.12160

17. Sagud M, Mihaljevic Peles A, Pivac N. Smoking in schizophrenia: Recent findings about an old problem. Curr Opin Psychiatry. 2019;32(5):402–408. doi:10.1097/YCO.0000000000000529

18. Davies NM, Holmes M V, Davey Smith G. Reading Mendelian randomisation studies: a guide, glossary, and checklist for clinicians. BMJ. 2018;362:k601. doi:10.1136/BMJ.K601

19. Davey Smith G, Ebrahim S. ‘Mendelian randomization’: can genetic epidemiology contribute to understanding environmental determinants of disease?*. Int J Epidemiol. 2003;32(1):1–22. doi:10.1093/ije/dyg070

20. Consortium SWG of the Pg, Ripke S, Walters JT, O’Donovan MC. Mapping genomic loci prioritises genes and implicates synaptic biology in schizophrenia. medRxiv. September 2020:2020.09.12.20192922. doi:10.1101/2020.09.12.20192922

21. Nelson CP, Goel A, Butterworth AS, et al. Association analyses based on false discovery rate implicate new loci for coronary artery disease. Nat Genet. 2017;49(9):1385–1391. doi:10.1038/ng.3913

22. Shah S, Henry A, Roselli C, et al. Genome-wide association and Mendelian randomisation analysis provide insights into the pathogenesis of heart failure. Nat Commun. 2020;11(1). doi:10.1038/s41467-019-13690-5

23. Evangelou E, Warren HR, Mosen-Ansorena D, et al. Genetic analysis of over 1 million people identifies 535 new loci associated with blood pressure traits. Nat Genet. 2018;50(10):1412–1425. doi:10.1038/s41588-018-0205-x

24. Nolte IM, Munoz ML, Tragante V, et al. Genetic loci associated with heart rate variability and their effects on cardiac disease risk. Nat Commun. 2017;8. doi:10.1038/ncomms15805

25. Arking DE, Pulit SL, Crotti L, et al. Genetic association study of QT interval highlights role for calcium signaling pathways in myocardial repolarization. Nat Genet. 2014;46(8):826–836. doi:10.1038/ng.3014

26. Verweij N, Benjamins J-W, Morley MP, et al. The Genetic Makeup of the Electrocardiogram. Cell Syst. 2020;11(3):229-238.e5. doi:10.1016/j.cels.2020.08.005

27. Liu M, Jiang Y, Wedow R, et al. Association studies of up to 1.2 million individuals yield new insights into the genetic etiology of tobacco and alcohol use. Nat Genet. January 2019:1. doi:10.1038/s41588-018-0307-5

28. Wootton R, Richmond R, Stuijfzand B, et al. Causal effects of lifetime smoking on risk for depression and schizophrenia: Evidence from a Mendelian randomisation study. Psychol Med. 2020;50(14):2435–2443. doi:10.1101/381301

29. Locke AE, Kahali B, Berndt SI, et al. Genetic studies of body mass index yield new insights for obesity biology. Nature. 2015;518(7538):197–206. doi:10.1038/nature14177

30. Doherty A, Smith-Byrne K, Ferreira T, et al. GWAS identifies 14 loci for device-measured physical activity and sleep duration. Nat Commun. 2018;9(1):5257. doi:10.1038/s41467-018-07743-4

31. Willer CJ, Schmidt EM, Sengupta S, et al. Discovery and refinement of loci associated with lipid levels. Nat Genet. 2013;45(11):1274–1285. doi:10.1038/ng.2797

32. Mahajan A, Taliun D, Thurner M, et al. Fine-mapping type 2 diabetes loci to single-variant resolution using high-density imputation and islet-specific epigenome maps. Nat Genet. 2018;50(11):1505–1513. doi:10.1038/s41588-018-0241-6

33. Finucane HK, Bulik-Sullivan B, Gusev A, et al. Partitioning heritability by functional annotation using genome-wide association summary statistics. Nat Genet. 2015;47(11):1228–1235. doi:10.1038/ng.3404

34. Bulik-Sullivan BK, Loh P-R, Finucane HK, et al. LD Score regression distinguishes confounding from polygenicity in genome-wide association studies. Nat Genet. 2015;47(3):291–295. doi:10.1038/ng.3211

35. Bowden J, Davey Smith G, Haycock PC, Burgess S. Consistent Estimation in Mendelian Randomization with Some Invalid Instruments Using a Weighted Median Estimator. Genet Epidemiol. 2016;40(4):304–314. doi:10.1002/gepi.21965

36. Hartwig FP, Smith GD, Bowden J. Robust inference in two-sample Mendelian randomisation via the zero modal pleiotropy assumption. Int J Epidemiol. 2017;46(6):1985–1998.

37. Bowden J, Davey Smith G, Burgess S. Mendelian randomization with invalid instruments: effect estimation and bias detection through Egger regression. Int J Epidemiol. 2015;44(2):512–525. doi:10.1093/ije/dyv080

38. Verbanck M, Chen CY, Neale B, Do R. Detection of widespread horizontal pleiotropy in causal relationships inferred from Mendelian randomization between complex traits and diseases. Nat Genet. 2018;50(5):693–698. doi:10.1038/s41588-018-0099-7

39. Zhu Z, Zheng Z, Zhang F, et al. Causal associations between risk factors and common diseases inferred from GWAS summary data. Nat Commun. 2018;9(1):224. doi:10.1038/s41467-017-02317-2

40. Hemani G, Tilling K, Davey Smith G. Orienting The Causal Relationship Between Imprecisely Measured Traits Using Genetic Instruments. PLOS Genet. 2017;13(11):e1007081. doi:10.1101/117101

41. Sanderson E. Multivariable Mendelian Randomization and Mediation. Cold Spring Harb Perspect Med. 2021;11(2):a038984. doi:10.1101/cshperspect.a038984

42. Sanderson E, Davey Smith G, Windmeijer F, Bowden J. An examination of multivariable Mendelian randomization in the single-sample and two-sample summary data settings. Int J Epidemiol. 2019;48(3):713–727. doi:10.1093/ije/dyy262

43. Sterne JAC, Smith GD, Cox DR. Sifting the evidence—what’s wrong with significance tests? BMJ. 2001;322(7280):226. doi:10.1136/bmj.322.7280.226

44. Sanderson E, Spiller W, Bowden J. Testing and Correcting for Weak and Pleiotropic Instruments in Two-Sample Multivariable Mendelian Randomisation. bioRxiv. April 2020:2020.04.02.021980. doi:10.1101/2020.04.02.021980

45. Pouget JG, Han B, Wu Y, et al. Cross-disorder analysis of schizophrenia and 19 immune-mediated diseases identifies shared genetic risk. Hum Mol Genet. 2019;28(20):3498–3513. doi:10.1093/hmg/ddz145

46. Vermeulen JM, Wootton RE, Treur JL, et al. Smoking and the risk for bipolar disorder: evidence from a bidirectional Mendelian randomisation study. Br J Psychiatry. September 2019:1–7. doi:10.1192/bjp.2019.202

47. Correll CU, Solmi M, Veronese N, et al. Prevalence, incidence and mortality from cardiovascular disease in patients with pooled and specific severe mental illness: a large-scale meta-analysis of 3,211,768 patients and 113,383,368 controls. World Psychiatry. 2017;16(2):163–180. doi:10.1002/wps.20420

48. Pillinger T, Osimo EF, de Marvao A, et al. Cardiac structure and function in patients with schizophrenia taking antipsychotic drugs: an MRI study. Transl Psychiatry. 2019;9(1). doi:10.1038/s41398-019-0502-x

49. Fitzgerald JL, Hay K, Sheridan J, Chadwick A, Burke A, Haqqani HM. Late Potentials and Early Repolarisation Are Associated With Serious Mental Illness and May Portend Increased Arrhythmic Risk. Hear Lung Circ. 2020;29(10):1476–1483. doi:10.1016/j.hlc.2020.02.012

50. Kameyama H, Sugimoto K, Inamura K, et al. Early repolarization pattern is associated with schizophrenia: A single center experience in Japan. medRxiv. July 2020:2020.07.16.20155838. doi:10.1101/2020.07.16.20155838

51. Derval N, Shah A, Jaïs P. Definition of early repolarization: A tug of war. Circulation. 2011;124(20):2185–2186. doi:10.1161/CIRCULATIONAHA.111.064063

52. Antonio Centurion O, Hocini M, Bourier F, et al. Early Repolarization Syndrome: Diagnostic and Therapeutic Approach. Front Cardiovasc Med | www.frontiersin.org. 2018;1:169. doi:10.3389/fcvm.2018.00169

53. Quintana DS, Westlye LT, Kaufmann T, et al. Reduced heart rate variability in schizophrenia and bipolar disorder compared to healthy controls. Acta Psychiatr Scand. 2016;133(1):44–52. doi:10.1111/acps.12498

54. Iwamoto Y, Kawanishi C, Kishida I, et al. Dose-dependent effect of antipsychotic drugs on autonomic nervous system activity in schizophrenia. BMC Psychiatry. 2012;12(1):1–6. doi:10.1186/1471-244X-12-199

55. Latvala A, Kuja-Halkola R, Rück C, et al. Association of resting heart rate and blood pressure in late adolescence with subsequent mental disorders: A longitudinal population study of more than 1 million men in Sweden. JAMA Psychiatry. 2016;73(12):1268–1275. doi:10.1001/jamapsychiatry.2016.2717

56. Cordes J, Bechdolf A, Engelke C, et al. Prevalence of metabolic syndrome in female and male patients at risk of psychosis. Schizophr Res. 2017;181:38–42. doi:10.1016/j.schres.2016.09.012

57. Burke GM, Genuardi M, Shappell H, D’Agostino RB, Magnani JW. Temporal Associations Between Smoking and Cardiovascular Disease, 1971 to 2006 (from the Framingham Heart Study). Am J Cardiol. 2017;120(10):1787–1791. doi:10.1016/j.amjcard.2017.07.087

58. Firth J, Solmi M, Wootton RE, et al. A meta-review of “lifestyle psychiatry”: the role of exercise, smoking, diet and sleep in the prevention and treatment of mental disorders. World Psychiatry. 2020;19(3):360–380. doi:10.1002/wps.20773

59. Martin J, Tilling K, Hubbard L, et al. Association of genetic risk for schizophrenia with nonparticipation over time in a population-based cohort study. Am J Epidemiol. 2016;183(12):1149–1158. doi:10.1093/aje/kww009

60. Tyrrell J, Zheng J, Beaumont R, et al. Genetic predictors of participation in optional components of UK Biobank. Nat Commun. 2021;12(1):1–13. doi:10.1038/s41467-021-21073-y

61. E S, S N, SP F, et al. Susceptibility Loci for Clinical Coronary Artery Disease and Subclinical Coronary Atherosclerosis Throughout the Life-Course. Circ Cardiovasc Genet. 2015;8(6):803–811. doi:10.1161/CIRCGENETICS.114.001071

62. Brumpton B, Sanderson E, Hartwig FP, et al. Within-family studies for Mendelian randomization: Avoiding dynastic, assortative mating, and population stratification biases. Nat Commun. July 2020:3519. doi:10.1101/602516

63. Nielsen RE, Banner J, Jensen SE. Cardiovascular disease in patients with severe mental illness. Nat Rev Cardiol. 2021;18(2):136–145. doi:10.1038/s41569-020-00463-7

