## Supplemental text for "Exploring the relationship between schizophrenia and cardiovascular disease: A genetic correlation and multivariable Mendelian randomization study"

**Content**

1. Justification for cardiovascular disease phenotypes	2-3
2. Data samples used and GWAS procedures	4-6
3. Sensitivity methods	7-10
4. Supplementary Figures 1-4 (leave-one-out analyses main findings)	11-14
5. Supplementary Figures 5-8 (scatterplots main findings)	15-16

### Justification for cardiovascular disease phenotypes

The cardiovascular disease (CVD) phenotypes that were included in our study were selected based on their relevance in relation to schizophrenia (based on clinical and epidemiological literature) as well as the sample size of the available GWAS. Only phenotypes that were both relevant and for which a large and powerful enough GWAS was available, were selected. Crucially, the specific phenotypes were, before data analysis started, pre-registered (<https://osf.io/fprew>; Study 2).

We selected *coronary artery disease (CAD)* and *heart failure (HF)*, as two commonly measured clinical endpoints known to be more prevalent in patients with schizophrenia compared to controls.^1,2^ Whereas CAD is a manifestation of a build-up ‘plaque’ in the coronary arteries (atherosclerosis), potentially leading to a myocardial infarction, HF has a more diverse aetiology. HF is considered a clinical syndrome of cardiac dysfunction, that often results from impairment of left ventricular function. Multiple cardiovascular and systemic disorders, including CAD, can be aetiological factors for the development of HF.^3^

In addition to the above mentioned clinical endpoints, we selected six important CVD risk markers. These include: *systolic blood pressure (SBP), diastolic blood pressure (DBP), heart rate variability, QT interval, early repolarization electrocardiogram (ECG) pattern and dilated cardiomyopathy ECG pattern*. SBP and DBP are well known and commonly measured markers of (poor) cardiovascular health, which are on average elevated in patients with schizophrenia compared to healthy controls.^4^ Heart rate variability is a general measure of variation in the cardiac cycle duration, which reflects the degree of cardiac vagal control***.*** A lower cardiac vagal control is associated with a higher risk of cardiac morbidity and mortality. Low heart rate variability is a prominent CVD marker reported in patients with schizophrenia*.*^5,6^ Lastly, our selection of CVD risk markers included three ECG patterns that are known to associate with a higher risk of cardiovascular disease or death. First, the QT interval, for which a particularly large GWAS was available.^7^ The QT interval is a fragment of the ECG that reflects myocardial repolarization. When the QT interval is prolongated, it increases the risk of ventricular arrhythmias and sudden cardiac death. The QT interval is often prolongated in patients with schizophrenia, which is thought to be (in part) due to the use of antipsychotic medication such as clozapine.^2,6^ The final two ECG related traits that we included are early repolarization pattern and dilated cardiomyopathy [atterm. These phenotypes both stem from a particularly large and extensive GWAS conducted to explore the complete ECG.^8^ The two main phenotypes that were focussed on in this GWAS, and which were followed up with extensive follow-up analyses (see Table 1 in the main paper), were early repolarization pattern and dilated cardiomyopathy pattern. While early repolarization is in some cases considered a normal ECG pattern, there is recent evidence suggesting that in other cases it can be a risk factor for idiopathic ventricular fibrillation and sudden cardiac death.^9,10^ Dilated cardiomyopathy occurs when the muscle of the heart weakens and becomes enlarged, resulting in decreased pumping function. Both early repolarization and dilated cardiomyopathy appear more frequently among patients with schizophrenia compared to healthy controls, in part due to the use of antipsychotic medication.

### GWAS data samples used

For schizophrenia, we requested the European only summary statistics from the latest and largest available GWAS of the Psychiatric Genomics Consortium (this study is currently available as a pre-print on medrxiv; 53,386 cases and 77,258 controls).^11^ For coronary artery disease, we employed the largest available GWAS which was based on participants of the UK Biobank study (71,602 cases and 260,875 controls).^12^ Coronary artery disease cases were based on the ‘SOFT phenotype’, which includes not only (fatal or nonfatal) myocardial infarction, but also the procedures percutaneous transluminal coronary angioplasty (PTCA) and coronary artery bypass grafting (CABG), as well as self-reported angina. For heart failure, we used the particularly large GWAS of the Heart Failure Molecular Epidemiology for Therapeutic Targets (HERMES) Consortium (47,309 cases and 930,014 controls).^3^ For blood pressure, we used data from a single GWAS (n = 757,601) that investigated several blood pressure related phenotypes.^13^ To get an detailed grasp of potential different blood pressure mechanisms, while also keeping the number of included variables (and thus multiple testing burden) down, we chose to include systolic blood pressure and diastolic blood pressure. For heart rate variability, there was only one, relatively large GWAS available.^14^ We chose to use the measure of heart rate variability expressed as the root mean square of the successive differences of inter beat intervals (RMSSD), as this is widely considered to be the most informative measure. When heart rate variability was the exposure in MR analyses, the estimates were based on the full sample size of n = 46,952, while for the other analyses (genetic correlations or when heart rate variability was the outcome) the estimates were based on a smaller sample of n = 26,523. This is due the fact that in the GWAS study, they only tested the genome-wide significant SNPs in a larger, follow-up sample. For QT interval, a particularly large GWAS (n = 103,331) was available.^7^ For early repolarization and dilated cardiomyopathy, we used data from one large GWAS that elegantly investigated the complete ECG spectrum across 500 unique time-points (n = 63,700), unprecedented for this type of measurement.^8^ From that spectrum, we selected the two time points that were followed-up and confirmed to be indicative of the early repolarization and dilated cardiomyopathy (see Table 1 in the main paper).

For the univariable MR analyses, we made sure there was no sample overlap between exposure and outcome, as this could lead to bias and type 1 error inflation.

*Multivariable analyses*

For smoking, we used one large GWAS sample of the GSCAN consortium (GWAS & Sequencing Consortium of Alcohol and Nicotine use), which investigated multiple stages of smoking behaviour.^15^ We only included the phenotype of smoking initiation. When smoking initiation was the exposure, we took estimates from the publication of Liu et al., based on the full sample size n = ~1.2 million, when smoking initiation was the outcome we took estimates based on the sample that excluded both UK Biobank and 23andMe (n = 249 171). We were not able to include the phenotypes cigarettes per day and smoking cessation as these were analysed in samples of (former) smokers only, meaning that any follow-up MR analyses with those phenotypes as exposures would have to be stratified on smoking status of the participants in the schizophrenia GWAS, which we could not do because we only had summary-level data available. As an alternative, we therefore added the phenotype ‘Lifetime smoking’, which was constructed in a large GWAS study of UK Biobank participants (n = 462,690) and reflects a composite measure of smoking initiation, years of smoking, smoking heaviness, and smoking cessation.^16^ For BMI, we used the largest GWAS available (n = 339,224), which included participants from 125 different studies.^17^ For physical activity we used data from the largest available GWAS, conducted on UK Biobank participants only (n = 91,105). The measure of physical activity was obtained through activity trackers worn by the participants over a period of 7 days.^18^ To capture lipid levels, we included total cholesterol and triglycerides from the largest available GWAS study (n = 188,577).^19^ Finally, we employed the largest available GWAS for type 2 diabetes (n = 74,124 cases and n = 824,006 controls).^20^

To prevent sample overlap between smoking initiation and the CVD outcome variables, we employed summary-level data of the GWAS of smoking initiation that excluded UK Biobank individuals. The GWAS for physical activity and lifetime smoking were conducted in a sample of only UK Biobank participants, which means that we were unable to add those in multivariable MR analyses were CAD, SBP, and DBP, early repolarization or dilated cardiomyopathy were the outcome (as those outcomes were mostly or completely based on UK Biobank participants as well).

*Genome-wide association study (GWAS) on being underweight and overweight*

GWAS analyses were conducted on being underweight and being overweight in UK Biobank participants of European descent.^21^ For this analysis, we used the BMI measurement with UK Biobank field ID 21001. For the underweight GWAS, we ran a GWAS of participants classified as underweight (with BMI < 25; N = 2,323) versus individuals classified as normal range (25 ≤ BMI ≤ 30; N = 148,708). For the overweight GWAS, we ran a GWAS of participants classified as overweight (with BMI > 30; N = 303,518) versus individuals classified as normal range (25 ≤ BMI ≤ 30; N = 148,708). We ran linear-mixed model (LMM) GWASs in fastGWA^22^ on 10,616,040 single nucleotide polymorphisms (SNPs) that were imputed using the Haplotype Reference Consortium.^21^ The LMM GWAS controls for cryptic relatedness and population stratification by including a genetic relatedness matrix (GRM) in the model. As an additional control for population stratification, we included the first 25 PCs derived from the GRM. We also control for sex and age. For details on the quality control procedures, principal component analysis (PCA) to identify subjects of European ancestry, PCA to capture ancestry differences within the European participants, and the construction of the GRM, see Abdellaoui et al (2021).^23^

### Sensitivity methods

#### Univariable analyses

We applied six additional MR approaches, besides IVW, as sensitivity methods for the univariable analyses. First, weighted median regression which can provide a consistent causal estimate even when up to 50% of the weight of the genetic instrument does not satisfy the IV assumptions.^24^ Second, we applied weighted mode regression. The estimate from weighted mode regression is consistent if the most frequent value among the causal effect estimates is contributed by valid genetic variants, even if the majority of instruments are invalid.^25^ Third, we applied MR-Egger regression, which can be used to explicitly test for horizontal pleiotropy.^26^ MR-Egger is similar to IVW regression, except that the intercept term is not fixed at zero. The intercept estimated by MR-Egger analysis can be interpreted as the average horizontally pleiotropic effect of the included genetic variants. Crucially, the MR-Egger method relies on two assumptions: the INstrument Strength Independent of Direct Effect (InSIDE) assumption and the NO Measurement Error (NOME) assumption. The InSIDE assumption states that the pleiotropic effects are not correlated with the instrument strength. The NOME assumption states that the variance of SNP-exposure association estimates is negligible. MR-Egger is particularly powerful as it can produce an unbiased estimate of the causal effect even if all SNPs show (some) horizontal pleiotropy. However, an important limitation is that MR-Egger has a markedly lower level of statistical power compared to IVW and other sensitivity methods like weighted median regression. Fourth, we used MR pleiotropy residual sum and outlier (MR-PRESSO) analysis.^27^ MR-PRESSO consists of three steps: testing for horizontal pleiotropy (MR-PRESSO global test), correcting for horizontal pleiotropy using outlier removal (MR-PRESSO outlier test) and evaluating significant differences in the causal effects estimated before and after outlier removal (MR-PRESSO distortion test). In order to use MR-PRESSO, 50% of the genetic variants included in the instrument are required to satisfy the IV assumptions. Fifth, we used the generalized summary data-based MR (GSMR) approach.^28^ The GSMR method provides more statistical power than other MR methods, because it uses very low levels of LD between the IVs. In addition, while other methods assume that the effects of the IVs on the exposure are estimated without error, GSMR considers the sampling variation in the estimated effects of the IVs. The GSMR methods also detects and removes IVs that have pleiotropic effects on both exposure and outcome with the HEIDI-outlier. Sixth, we applied Steiger filtering to correct for reverse causality.^29^ With this method, SNPs were identified and then excluded if they explained a larger amount of variance in the outcome, compared to the exposure. If there is an actual causal effect from the exposure to the outcome, the SNPs should be more predictive of the exposure.

Apart from the above-mentioned formal MR methods, we also computed the Q, F and I^2^ statistics for all univariable analyses. To assess heterogeneity between individual SNP estimates, which might indicate violations of IV assumptions, we used the Cochran’s Q statistic. The Cochran’s Q statistic was computed for both the IVW and Egger estimates. The F statistic can be used to detect weak instruments. When bias increases, the F statistic decreases. Sufficient instrument strength is defined as a F statistic above 10. We used an adaptation of the I^2^ statistic known from meta-analysis to assess the NOME assumption for MR-Egger, also called I^2^_GX_.^30^ It indicates possible bias from the NOME assumption. If I^2^_GX_ is below 0.6, the MR-egger results are considered unreliable. In this case, they were not reported. A I^2^_GX_ higher than 0.9 indicates bias due to violation of the NOME assumption is not likely. When the I^2^_GX_ = 0.6 – 0.9, there is a possibility of bias caused by violation of the NOME assumption. Simulation extrapolation (SIMEX) was used to correct the MR-Egger estimates. Lastly, to examine whether the analyses were driven or biased by a single SNP, we conducted a leave-one-out analysis, performing IVW regression analyses in which we consecutively left one SNP out each time.

#### Multivariable analyses

For the multivariable MR analysis, we applied three sensitivity methods. First, we used the Sanderson–Windmeijer conditional F statistic to detect weak instruments, with a F statistic of more than 10 indicating sufficient instrument strength.^31^ Second, we used an adaptation of the Cochran’s Q statistic to detect heterogeneity among the SNPs included for analysis. Third, we applied the multivariable MR-Egger to test if horizontal pleiotropy was present, which is slightly different than the univariable MR-Egger method. In univariable MR-Egger, the SNPs are orientated so that the direction of the association with the exposure is either positive or negative for all SNPs. In multivariable MR-Egger this is not possible, so we repeated the analysis with different orientations to ensure reliable results. We reported the results orientated with respect to the exposure of interest, and observed if pleiotropy was absent in each of the different orientations.

*References*

1.	De Hert M, Detraux J, Vancampfort D. The intriguing relationship between coronary heart disease and mental disorders. *Dialogues Clin Neurosci*. 2018;20(1):31-40. http://www.ncbi.nlm.nih.gov/pubmed/29946209. Accessed February 5, 2019.

2.	Koponen H, Alaräisänen A, Saari K, et al. Schizophrenia and sudden cardiac death—A review. *Nord J Psychiatry*. 2008;62(5):342-345. doi:10.1080/08039480801959323

3.	Shah S, Henry A, Roselli C, et al. Genome-wide association and Mendelian randomisation analysis provide insights into the pathogenesis of heart failure. *Nat Commun*. 2020;11(1). doi:10.1038/s41467-019-13690-5

4.	Mitchell AJ, Vancampfort D, De Herdt A, Yu W, De Hert M. Is the prevalence of metabolic syndrome and metabolic abnormalities increased in early schizophrenia? a comparative meta-analysis of first episode, untreated and treated patients. *Schizophr Bull*. 2013;39(2):295-305. doi:10.1093/schbul/sbs082

5.	Quintana DS, Westlye LT, Kaufmann T, et al. Reduced heart rate variability in schizophrenia and bipolar disorder compared to healthy controls. *Acta Psychiatr Scand*. 2016;133(1):44-52. doi:10.1111/acps.12498

6.	Cohen H, Loewenthal U, Matar M, Kotler M. Association of autonomic dysfunction and clozapine: Heart rate variability and risk for sudden death in patients with schizophrenia on long-term psychotropic medication. *Br J Psychiatry*. 2001;179(2):167-171. doi:10.1192/BJP.179.2.167

7.	Arking DE, Pulit SL, Crotti L, et al. Genetic association study of QT interval highlights role for calcium signaling pathways in myocardial repolarization. *Nat Genet*. 2014;46(8):826-836. doi:10.1038/ng.3014

8.	Verweij N, Benjamins J-W, Morley MP, et al. The Genetic Makeup of the Electrocardiogram. *Cell Syst*. 2020;11(3):229-238.e5. doi:10.1016/j.cels.2020.08.005

9.	Derval N, Shah A, Jaïs P. Definition of early repolarization: A tug of war. *Circulation*. 2011;124(20):2185-2186. doi:10.1161/CIRCULATIONAHA.111.064063

10.	Antonio Centurion O, Hocini M, Bourier F, et al. Early Repolarization Syndrome: Diagnostic and Therapeutic Approach. *Front Cardiovasc Med | www.frontiersin.org*. 2018;1:169. doi:10.3389/fcvm.2018.00169

11.	Consortium SWG of the PG, Ripke S, Walters JT, O’Donovan MC. Mapping genomic loci prioritises genes and implicates synaptic biology in schizophrenia. *medRxiv*. September 2020:2020.09.12.20192922. doi:10.1101/2020.09.12.20192922

12.	Nelson CP, Goel A, Butterworth AS, et al. Association analyses based on false discovery rate implicate new loci for coronary artery disease. *Nat Genet*. 2017;49(9):1385-1391. doi:10.1038/ng.3913

13.	Evangelou E, Warren HR, Mosen-Ansorena D, et al. Genetic analysis of over 1 million people identifies 535 new loci associated with blood pressure traits. *Nat Genet*. 2018;50(10):1412-1425. doi:10.1038/s41588-018-0205-x

14.	Nolte IM, Munoz ML, Tragante V, et al. Genetic loci associated with heart rate variability and their effects on cardiac disease risk. *Nat Commun*. 2017;8. doi:10.1038/ncomms15805

15.	Liu M, Jiang Y, Wedow R, et al. Association studies of up to 1.2 million individuals yield new insights into the genetic etiology of tobacco and alcohol use. *Nat Genet*. January 2019:1. doi:10.1038/s41588-018-0307-5

16.	Wootton R, Richmond R, Stuijfzand B, et al. Causal effects of lifetime smoking on risk for depression and schizophrenia: Evidence from a Mendelian randomisation study. *Psychol Med*. 2020;50(14):2435-2443. doi:10.1101/381301

17.	Locke AE, Kahali B, Berndt SI, et al. Genetic studies of body mass index yield new insights for obesity biology. *Nature*. 2015;518(7538):197-206. doi:10.1038/nature14177

18.	Doherty A, Smith-Byrne K, Ferreira T, et al. GWAS identifies 14 loci for device-measured physical activity and sleep duration. *Nat Commun*. 2018;9(1):5257. doi:10.1038/s41467-018-07743-4

19.	Willer CJ, Schmidt EM, Sengupta S, et al. Discovery and refinement of loci associated with lipid levels. *Nat Genet*. 2013;45(11):1274-1285. doi:10.1038/ng.2797

20.	Mahajan A, Taliun D, Thurner M, et al. Fine-mapping type 2 diabetes loci to single-variant resolution using high-density imputation and islet-specific epigenome maps. *Nat Genet*. 2018;50(11):1505-1513. doi:10.1038/s41588-018-0241-6

21.	C B, C F, D P, et al. The UK Biobank resource with deep phenotyping and genomic data. *Nature*. 2018;562(7726):203-209. doi:10.1038/S41586-018-0579-Z

22.	Jiang L, Zheng Z, Qi T, et al. A resource-efficient tool for mixed model association analysis of large-scale data. *Nat Genet*. 2019;51(12):1749-1755. doi:10.1038/s41588-019-0530-8

23.	Abdellaoui A, Verweij KJH, Nivard MG. Geographic Confounding in Genome-Wide Association Studies. *bioRxiv*. March 2021:2021.03.18.435971. doi:10.1101/2021.03.18.435971

24.	Bowden J, Davey Smith G, Haycock PC, Burgess S. Consistent Estimation in Mendelian Randomization with Some Invalid Instruments Using a Weighted Median Estimator. *Genet Epidemiol*. 2016;40(4):304-314. doi:10.1002/gepi.21965

25.	Hartwig FP, Smith GD, Bowden J. Robust inference in two-sample Mendelian randomisation via the zero modal pleiotropy assumption. *Int J Epidemiol*. 2017;46(6):1985-1998.

26.	Bowden J, Davey Smith G, Burgess S. Mendelian randomization with invalid instruments: effect estimation and bias detection through Egger regression. *Int J Epidemiol*. 2015;44(2):512-525. doi:10.1093/ije/dyv080

27.	Verbanck M, Chen CY, Neale B, Do R. Detection of widespread horizontal pleiotropy in causal relationships inferred from Mendelian randomization between complex traits and diseases. *Nat Genet*. 2018;50(5):693-698. doi:10.1038/s41588-018-0099-7

28.	Zhu Z, Zheng Z, Zhang F, et al. Causal associations between risk factors and common diseases inferred from GWAS summary data. *Nat Commun*. 2018;9(1):224. doi:10.1038/s41467-017-02317-2

29.	Hemani G, Tilling K, Davey Smith G. Orienting The Causal Relationship Between Imprecisely Measured Traits Using Genetic Instruments. *PLOS Genet*. 2017;13(11):e1007081. doi:10.1101/117101

30.	Bowden J, Del Greco M. F, Minelli C, Davey Smith G, Sheehan NA, Thompson JR. Assessing the suitability of summary data for two-sample Mendelian randomization analyses using MR-Egger regression: the role of the I2 statistic. *Int J Epidemiol*. 2016;45(6):dyw220. doi:10.1093/ije/dyw220

31.	Sanderson E, Spiller W, Bowden J. Testing and Correcting for Weak and Pleiotropic Instruments in Two-Sample Multivariable Mendelian Randomisation. *bioRxiv*. April 2020:2020.04.02.021980. doi:10.1101/2020.04.02.021980

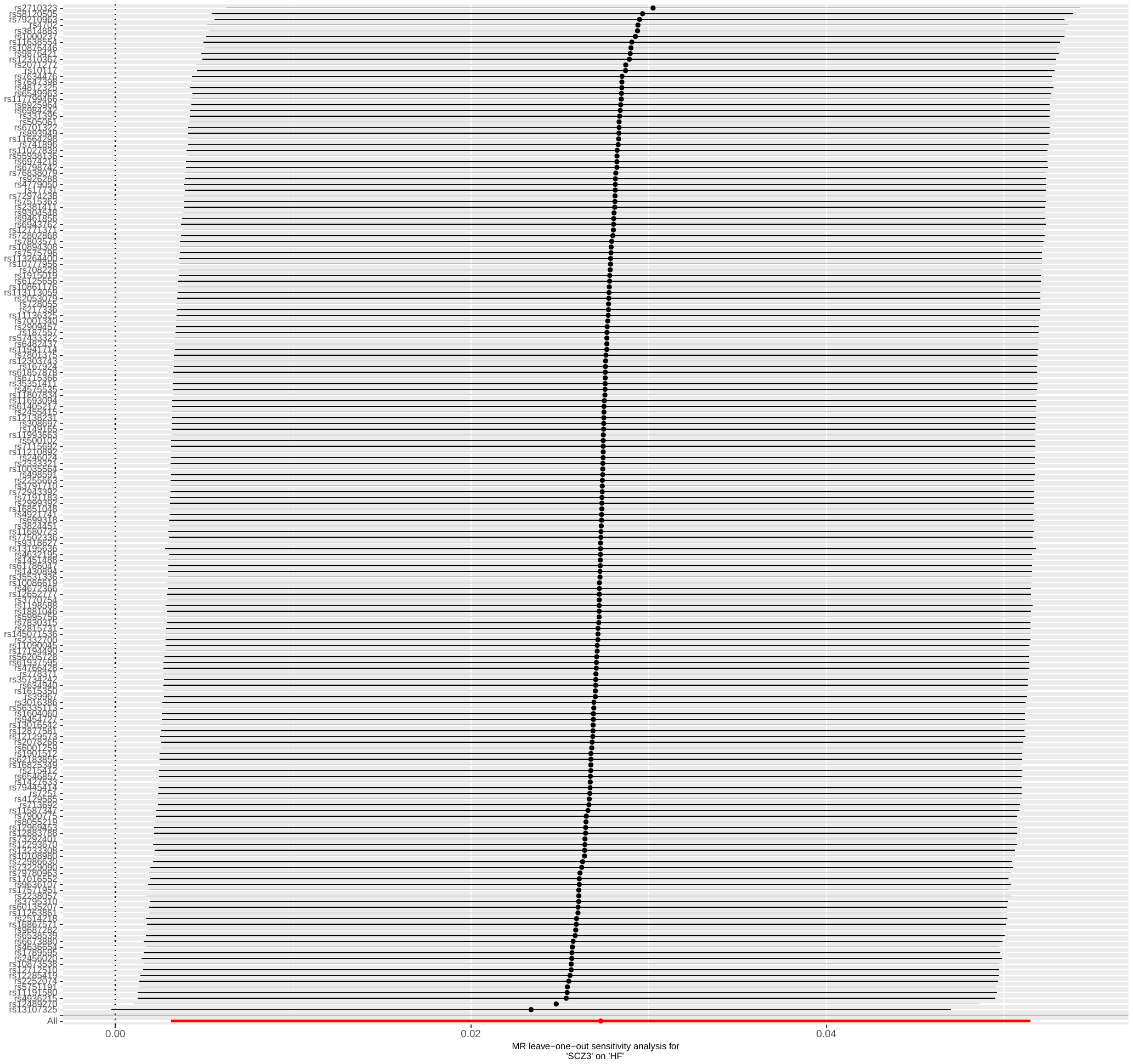

#### **Figure S1**. Leave-one-out IVW regression analyses of liability to schizophrenia on heart failure risk

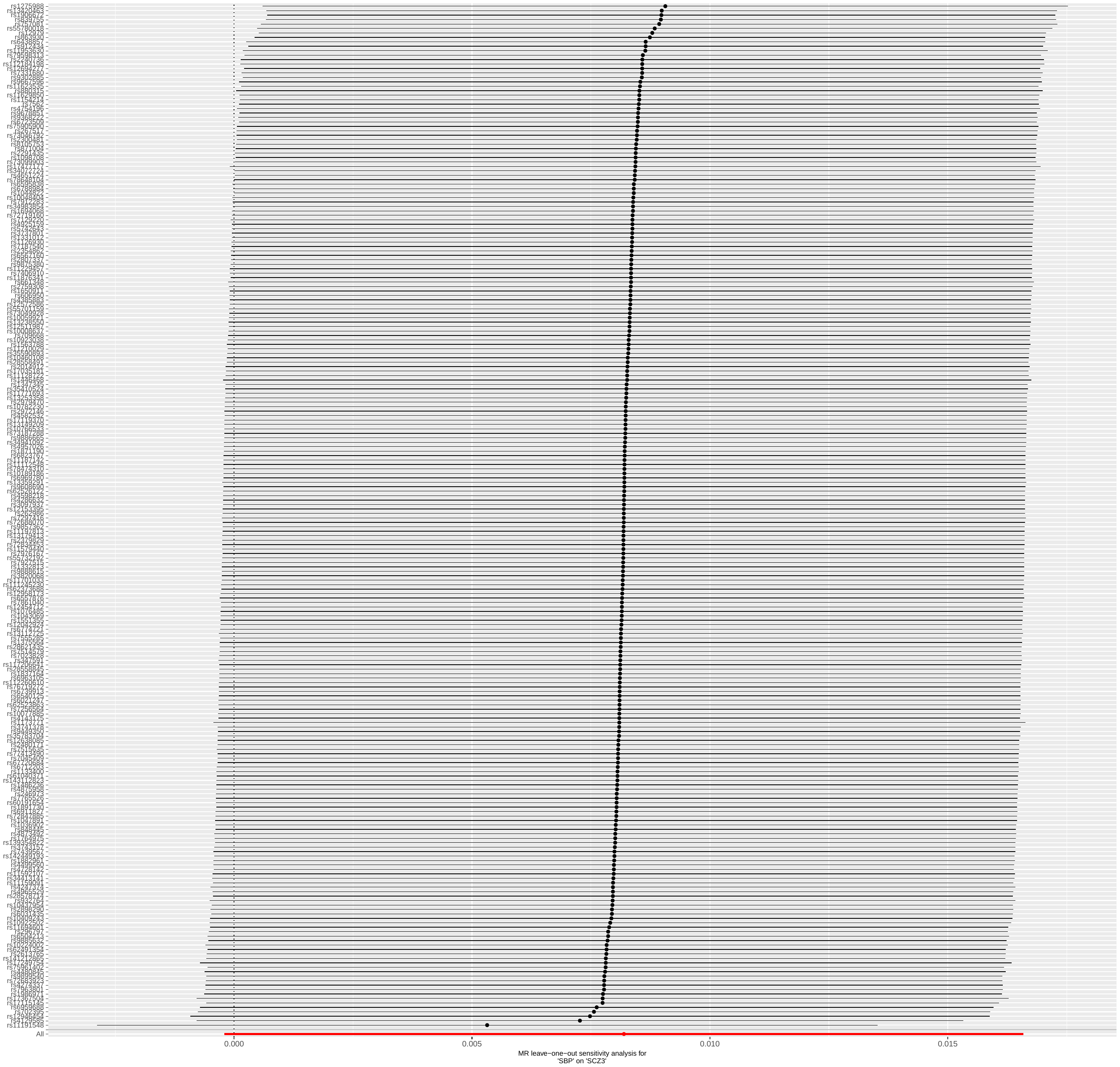

#### **Figure S2**. Leave-one-out IVW regression analyses of liability to systolic blood pressure on schizophrenia risk

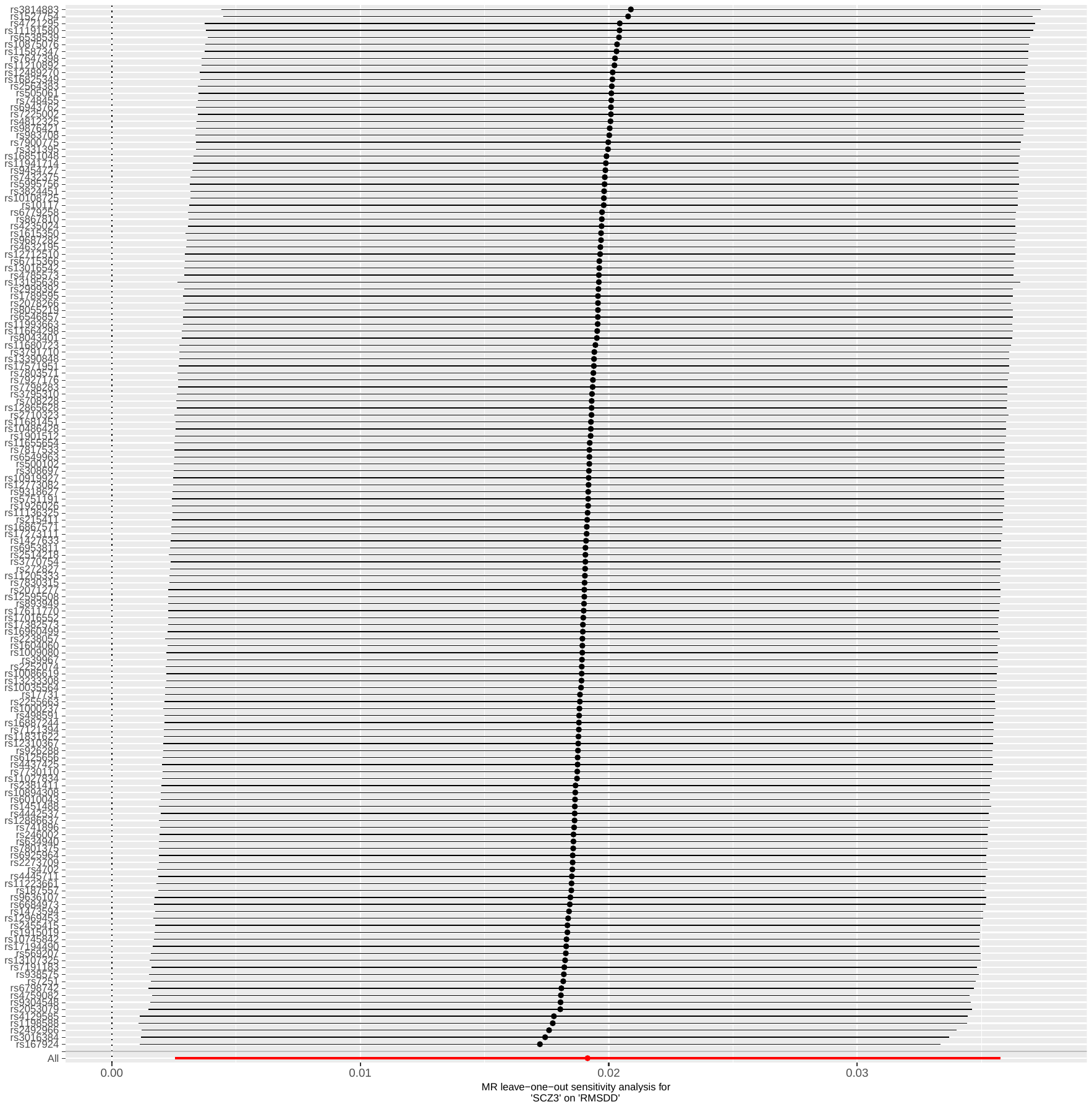

#### **Figure S3**. Leave-one-out IVW regression analyses of liability to schizophrenia on heart rate variability

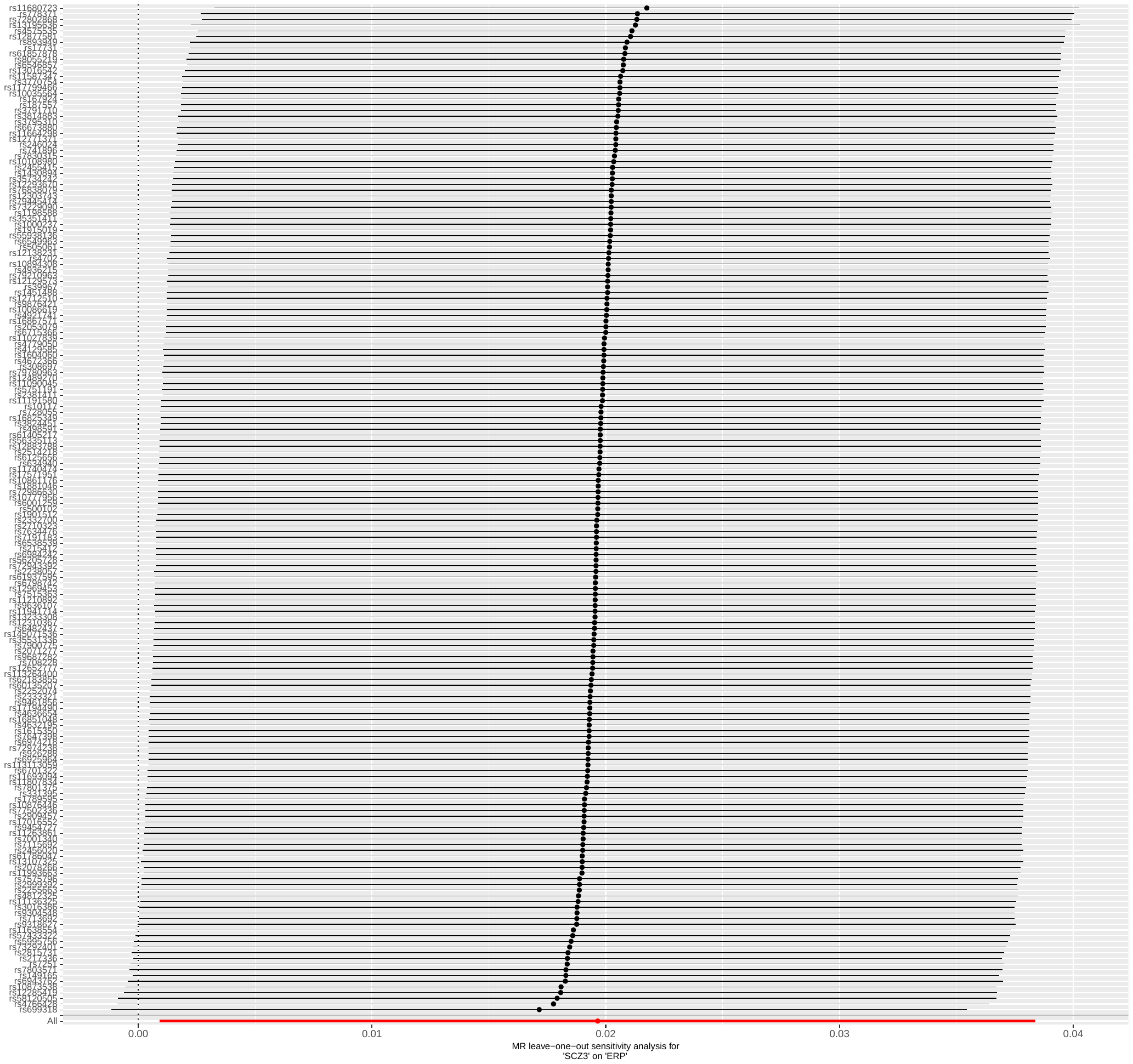

#### **Figure S4**. Leave-one-out IVW regression analyses of liability to schizophrenia on early repolarization risk

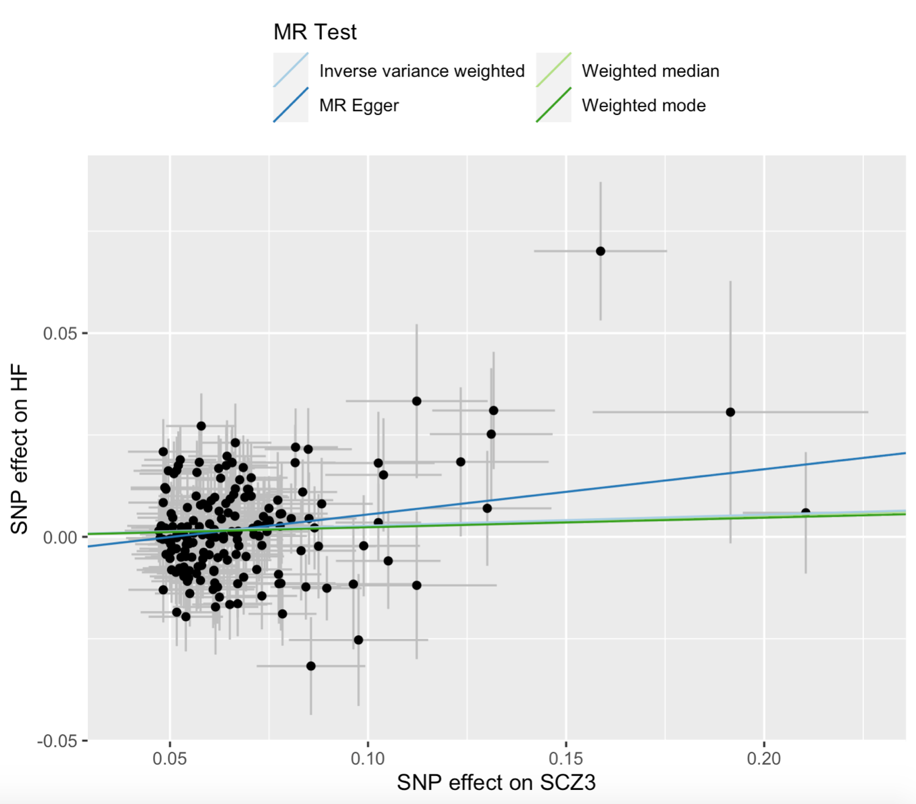

#### **Figure S5**. Scatter plot of the univariable MR analysis of the liability to schizophrenia on heart failure risk

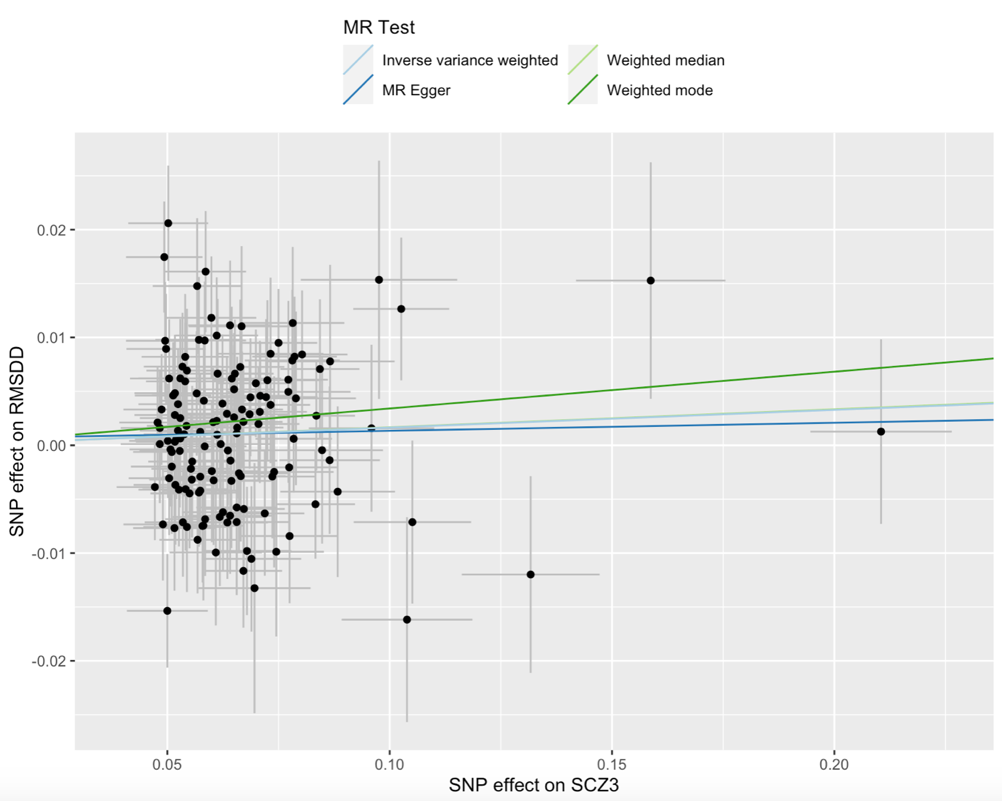

#### **Figure S6**. Scatter plot of the univariable MR analysis of the liability to schizophrenia on heart rate variability

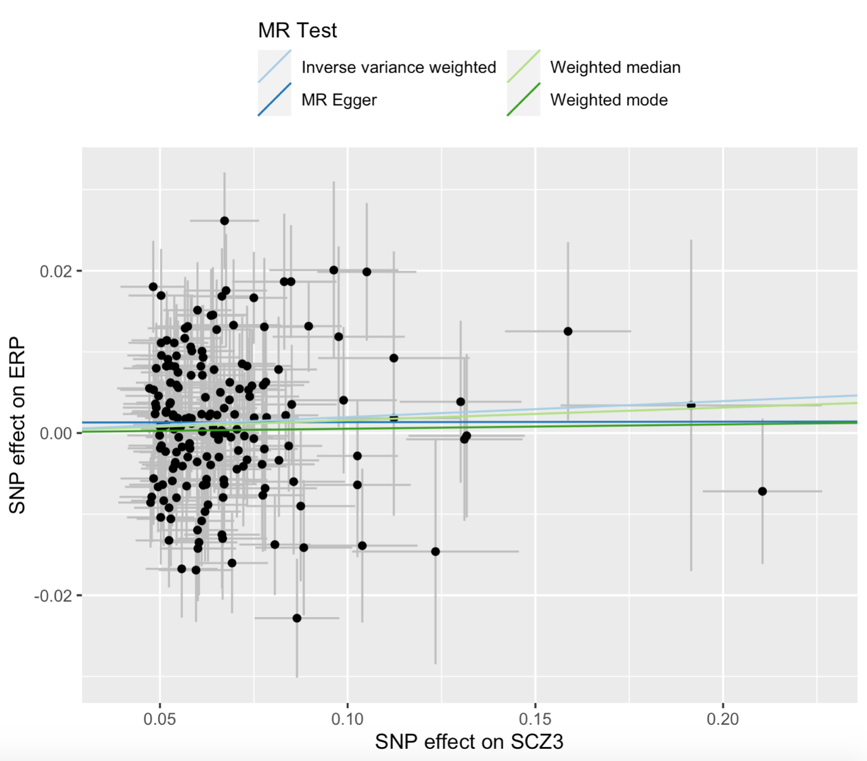

#### **Figure S7**. Scatter plot of the univariable MR analysis of the liability to schizophrenia on early repolarization risk

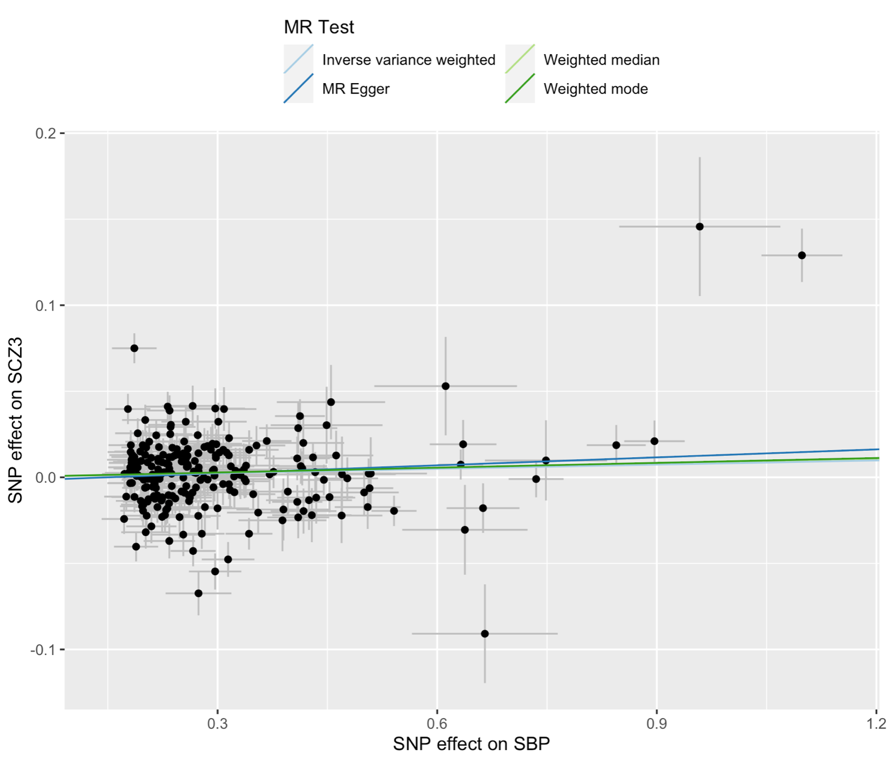

#### **Figure S8**. Scatter plot of the univariable MR analysis of the liability to systolic blood pressure on schizophrenia risk
